# Spatial biomarker-driven deep learning model via digital pathology predicts response to PI3K inhibitor buparlisib in head and neck squamous cell carcinoma

**DOI:** 10.1101/2025.10.09.25337502

**Authors:** Antoine Desilets, Minh Tri Le, Justin Lucas, Orit Matcovitch-Natan, Amit Bart, Avi Laniado, Meir Azulay, Ettai Markovits, Jennifer Kaplan Kerner, Amit Gutwillig, Hadar Yehezkeli, Lisa F. Licitra, Sunny Lu, Kevin Dreyer, Ying Pan, Nanhai He, Archie Tse, Sandrine Faivre, Denis Soulières

**Author notes:** **Corresponding author:** Antoine Desilets, MD, MSc; Hematology-Oncology Service, Department of Medicine; Centre hospitalier de l’Université de Montréal (CHUM), 1000, rue Saint-Denis, Montreal, QC, Canada.

## Abstract

**Purpose:** Buparlisib, a pan-class I PI3K inhibitor, combined with paclitaxel, demonstrated improved survival in the BERIL-1 trial for patients with recurrent/metastatic (R/M) head and neck squamous cell carcinoma (HNSCC). However, predictive biomarkers of benefit remain undefined. We evaluated spatial biomarkers derived from hematoxylin and eosin (H&E) images using artificial intelligence (AI)–based digital pathology.

**Materials and Methods:** Whole-slide H&E images (n=144) from BERIL-1 were analyzed using a deep learning model trained to segment tissue compartments and classify individual cell phenotypes. Three prospectively defined spatial features were evaluated: (1) tumor-infiltrating lymphocyte (TIL) density in the tumor area; (2) tumor microenvironment (TME) heterogeneity; and (3) granulocyte fraction in the tumor invasive margin (TIM). Cox proportional hazards model was used to evaluate biomarker–treatment interactions, with patients stratified by biomarker status.

**Results:** High TIL density (>10%) defined by deep learning–derived analysis of H&E was associated with a significantly improved overall survival with buparlisib versus placebo (HR□=□0.25; 95% CI, 0.01–0.64; p = 0.002), as were high TME heterogeneity (HR□=□0.47; 95% CI, 0.27–0.80; p = 0.005) and granulocyte enrichment in the TIM (HR = 0.51; p = 0.014); in a within-arm proximity analysis, higher granulocyte–tumor cell proximity correlated with improved OS on buparlisib (HR = 0.32; p < 0.001). AI-derived spatial metrics outperformed CD3 immunohistochemistry staining in stratifying survival outcomes. In patients with oropharyngeal tumors, human papillomavirus-positive cases were more frequent among those with high TILs.

**Conclusions:** Spatial features extracted from standard H&E slides using AI-driven digital pathology can predict OS benefit from buparlisib in R/M HNSCC. These cost-effective and scalable biomarkers support image-based patient selection strategies and are being prospectively evaluated in the ongoing BURAN phase 3 trial.

## Introduction

Recurrent or metastatic (R/M) head and neck squamous cell carcinoma (HNSCC) is a highly aggressive malignancy with poor survival outcomes. Despite advances with the advent of immune checkpoint inhibitors (ICI)^1,2^ and targeted agents^3-5^, the majority of patients eventually experience disease progression, underscoring the need for more effective and personalized treatment strategies. Among emerging therapeutic approaches, phosphoinositide 3-kinase (PI3K) inhibitors have shown promise^6,7^; however, clinical benefit has been heterogeneous across cohorts^8-10^. The lack of validated predictors of response has hindered efforts to identify patients most likely to benefit from PI3K inhibition, complicating the adoption of biomarker-guided strategies.

The BERIL-1 clinical trial demonstrated an objective response rate (ORR), progression-free survival, and overall survival (OS) benefit with the addition of buparlisib, a pan-class I PI3K inhibitor, to paclitaxel in patients with R/M HNSCC who had progressed on platinum-based chemotherapy^7^. Translational analyses from BERIL-1 identified several molecular and immunologic features associated with improved response, including *PIK3CA* and *TP53* mutations, high tumor mutational burden (TMB), and elevated baseline immune infiltration as measured by CD8+ T cells and tumor-infiltrating lymphocytes (TILs)^8,9^. However, these conventional biomarkers – typically applied in a binary manner using predefined thresholds – fail to capture the spatial architecture and complexity of the tumor microenvironment (TME), which is increasingly recognized as a key modulator of therapeutic response.

Conventional diagnostic approaches in pathology, such as immunohistochemistry (IHC) and bulk DNA or RNA sequencing via next-generation sequencing (NGS), offer limited insight into the spatial heterogeneity and cellular architecture of the tumor microenvironment (TME). IHC provides low spatial resolution and is subject to interpretive variability, while NGS can be costly, technically complex, and not universally accessible. In contrast, hematoxylin and eosin (H&E) staining is inexpensive, routinely performed, and widely available across pathology laboratories. Despite its ubiquity, H&E remains underutilized as a source of quantitative biomarker information, largely due to the historical lack of tools for extracting interpretable spatial features in a high-throughput and reproducible manner. Recent advances in artificial intelligence (AI)-driven digital pathology now enable the analysis of whole-slide H&E images to identify clinically meaningful patterns – such as immune infiltration, cellular heterogeneity, and stromal organization – with high spatial resolution. These features offer critical context for understanding the TME contexture that may influence drug susceptibility and resistance.

In this study, we applied a deep learning pipeline to digitized H&E slides from BERIL-1 to evaluate the prognostic and predictive value of spatial biomarkers. We hypothesized that AI-based spatial annotation of H&E images could identify tumor microenvironmental features – such as TIL density, TME heterogeneity, and granulocyte infiltration - that correlate with response to buparlisib. These features have biological plausibility: PI3K signaling is a central regulator of immune evasion, stromal remodeling, and myeloid cell recruitment, while chemotherapy can further modulate immune–stromal interactions. Thus, immunological and spatial features of the TME may directly influence sensitivity to combined PI3K inhibition and paclitaxel. Our goal was to explore whether digital H&E analysis could provide a cost-effective, scalable tool to refine patient selection in future PI3K inhibitor trials.

## Methods

### Study design

This retrospective exploratory study analyzed archival H&E-stained tumor slides from patients enrolled in the BERIL-1 trial, a multicenter, randomized, double-blind, placebo-controlled phase II study in adults with histologically or cytologically confirmed R/M HNSCC who had progressed following platinum-based chemotherapy. Patients were randomized to receive intravenous paclitaxel (80 mg/m^2^ on days 1, 8, 15, and 22 of each 28-day cycle) in combination with either daily oral buparlisib (100 mg) or placebo. The primary endpoint was PFS, with OS and ORR as secondary endpoints. Radiological responses were assessed by Response Evaluation Criteria in Solid Tumors (RECIST) version 1.1. Tumor samples were obtained prior to treatment initiation and originated from either primary or metastatic sites. All patients provided written informed consent prior to study participation. The trial was conducted in accordance with the Declaration of Helsinki, approved by IRBs at all participating institutions and registered at ClinicalTrials.gov (NCT01852292).

### Tumor samples and image digitization

Formalin-fixed, paraffin-embedded (FFPE) sections from tumor samples at baseline were evaluated to ensure sufficient tumor content and staining quality. Only slides with sufficient tumor content, intact tissue morphology, and acceptable staining quality were digitized for analysis. Slides that failed QC were excluded due to issues such as absence of tumor cells (e.g., slides containing only blood or benign tissue) or poor technical quality (e.g., degraded or poorly stained tissue). Of the 156 screened slides, 12 (8%) were excluded following this QC process (see **Supplemental Table S1**). Conventional TIL IHC scoring was performed using CD3 immunostaining on a single whole-tissue section, with lymphocyte density assessed across intratumoral and stromal compartments by visual estimation. Human papillomavirus (HPV) status was assessed in all patients via p16 immunohistochemistry on FFPE tumor samples, regardless of primary tumor site, and correlated with TIL infiltration. Slides were scanned at 40× magnification using a high-resolution whole-slide imaging (WSI) system. The digitized images underwent a standardized quality control review for staining consistency, cellular integrity, and completeness of tumor tissue architecture. Slides were excluded if they contained only blood or benign tissue, if necrosis predominated, or if severe staining artifacts were present. Following this QC process, 144 slides were retained for analysis out of 156 screened (12 excluded, ∼8%). Full QC details, including exclusion categories, are provided in **Supplemental Table S1**. This curated dataset enabled the generation of spatially annotated maps of the tumor and immune microenvironment, forming the basis for subsequent correlation with clinical endpoints.

### Deep learning model development using digital pathology

Digitized WSIs were analyzed using a deep learning–based digital pathology platform designed to extract spatially resolved features of the TME at a single-cell resolution. The pipeline consisted of various convolution neural networks (CNNs) for different tasks. The two core modules are: (1) a spatial segmentation model to delineate histologic compartments, and (2) a single-cell classification model to characterize the phenotypic composition of the TME, encompassing principal immune and stromal cell types. The spatial segmentation model partitioned the tissue into tumor center (TC), tumor invasive margin (TIM), adjacent TME (aTME), and outer TME (oTME), using a pre-defined 60 µm inner and outer boundary relative to the tumor-stroma interface. These regions were color-coded and overlaid on the H&E slides to enable interpretable visualization of the TME structure (**Figure 1A**). Cell-type annotations derived from the classification model were rendered at single-cell resolution and superimposed on representative high-resolution zones to visualize tumor cells, lymphocytes, plasma cells, fibroblasts, endothelial cells, and granulocytes (**Figure 1B**). The deep learning models used for cell classification and spatial segmentation were first trained on external histopathology datasets and subsequently adapted and internally validated on a subset of BERIL-1 cases with expert annotations and reviewed by pathologists, ensuring generalizability while maintaining study-specific relevance.

**Figure 1.**
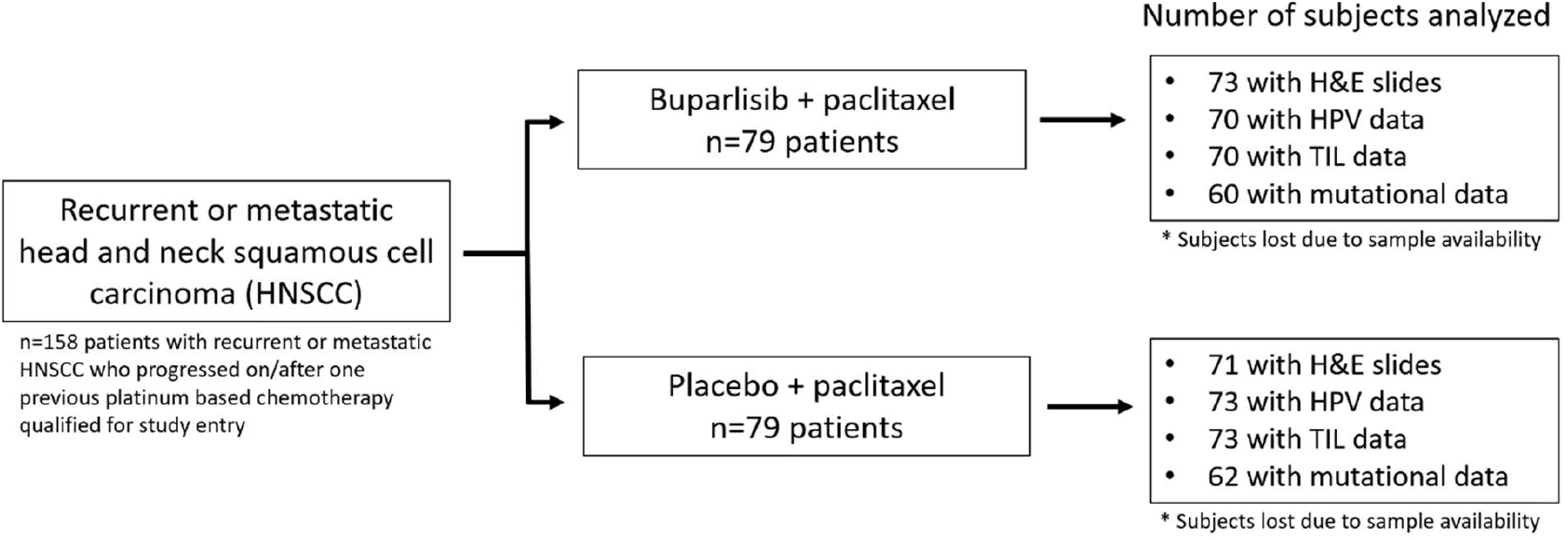
BERIL-1 trial design and translational analysis sample size. Overview of the BERIL-1 phase II clinical trial, which enrolled n=158 patients with recurrent or metastatic head and neck squamous cell carcinoma (HNSCC) progressing after prior platinum-based chemotherapy. Patients were randomized to receive either buparlisib plus paclitaxel or placebo plus paclitaxel (n=79 per arm). Translational analyses were conducted on available tissue samples. In the buparlisib arm, 73 patients had H&E slides for digital pathology analysis, 70 patients had HPV and TIL data, and 60 had mutational data from targeted DNA next-generation sequencing (NGS). In the placebo arm, 71 patients had H&E slides for digital pathology analysis, 73 had HPV data, 73 had TIL data, and 62 had mutational data. A total of 76 patients from each arm were included in the H&E-based digital pathology analysis based on tissue availability. H&E: hematoxylin and eosin; HPV: human papillomavirus; TIL: tumor-infiltrating lymphocytes

### Evaluation of single-cell classification and spatial segmentation models

The single-cell classification model applied deep convolutional neural networks trained on expert-labeled data to assign phenotypic identities to individual cells. Targeted cell types included tumor cells, lymphocytes, fibroblasts, plasma cells, granulocytes, and endothelial cells. Model performance was evaluated by comparing predictions with reference annotations provided by expert annotators and reviewed by board-certified pathologists (co-author JKK) in head and neck cancer. Although not fully blinded, comparisons were repeated across multiple runs, and a final holistic evaluation was performed to ensure accuracy across whole-slide images. In parallel, the spatial segmentation model was assessed across five distinct histological regions – tumor, stroma, necrosis, nerve, and other non-neoplastic tissue – by computing concordance between predicted and reference labels at the region level. Only cells with well-defined morphology were included in the ground truth dataset to ensure robust assessment. All performance metrics were independently reviewed and validated by a board-certified pathologist (JKK) to ensure interpretative consistency and accuracy.

### Spatial biomarker derivation and quantification

Three spatial biomarkers were prospectively defined and extracted from annotated H&E slides for each patient. These were selected from a larger pool of spatial features (∼3000 calculated) based on predefined biological and clinical rationale: (1) the density of TILs within the tumor area – defined as the tumor center and tumor invasive margin, excluding stromal regions (TC + TIM); (2) cellular heterogeneity of the TME, quantified using the Shannon entropy index across all classified cell phenotypes – a validated non-parametric index of ecological diversity; and (3) the proportion of granulocytes within the tumor invasive margin (TIM), representative of localized innate immune infiltration. Spatial features were computed using AI-based single-cell classification and region-specific overlays. TME heterogeneity and granulocyte fraction were each dichotomized as high or low according to the median value across the cohort. For TILs, a pre-specified threshold of 10% was used based on prior biological and clinical rationale^8,9^. All biomarker metrics were derived independently of clinical outcomes and later evaluated for association with OS in exploratory analyses. In a secondary proximity analysis, we also quantified a granulocyte–tumor cell proximity metric, defined as the proportion of granulocytes located within 60 µm of tumor cells in the tumor area (TC + TIM) using the same region overlays; this metric was used for within-arm associations.

### Statistical analysis

The primary clinical endpoint for this retrospective analysis was OS, defined as the time from randomization to death from any cause. OS distributions were estimated using the Kaplan-Meier method and compared between the treatment arms within each of the biomarker-defined subgroups using log-rank tests. Cox proportional hazards models were used to estimate hazard ratios (HRs) between the treatment arms and corresponding 95% confidence intervals (CIs) within each biomarker-defined subgroup. All biomarker analyses were exploratory; two-sided p-values < 0.05 were considered nominally significant without adjustment for multiplicity. Given the exploratory nature of this analysis, no adjustments were made for multiple comparisons. All analyses were conducted in R version 4.2.2 (R Foundation for Statistical Computing, Vienna, Austria). Primary analyses focused on predictive effects, estimating the treatment effect of buparlisib versus placebo within each biomarker-defined subgroup (high/low). Prognostic, within-arm high-vs-low comparisons were performed only as secondary analyses (e.g., the granulocyte–tumor-cell proximity metric).

## Results

### Survival outcomes and biomarker landscape of BERIL-1

Treatment with buparlisib in the BERIL-1 trial was associated with improved survival outcomes compared to placebo, including a median PFS of 4.6 vs. 3.5 months (HR = 0.65, nominal one-sided p = 0.011) and a median OS of 10.4 vs. 6.5 months (HR=0.72, nominal one-sided *p*=0.041). These efficacy results reflect the statistical approach of the original BERIL-1 trial, which used one-sided testing for primary and secondary endpoints. The objective response rate was also higher in the buparlisib arm (39% vs. 14%; nominal one-sided *p*<0.001), supporting the clinical benefit of PI3K inhibition in this population^7^. Prior translational analyses demonstrated a high frequency of oncogenic alterations - most commonly in *TP53, NOTCH1*, and *PIK3CA*. While genomic and IHC-based immune biomarkers, such as TILs and CD8+ cell infiltration, were broadly prevalent, they did not fully explain differential treatment benefit, prompting further evaluation of spatial biomarkers using digital pathology^8,9^. Unless otherwise specified, p-values for biomarker-defined subgroup analyses are reported as two-sided.

Among the 158 patients enrolled, 144 had available baseline FFPE tumor samples suitable for digital pathology analysis, enabling spatial biomarker extraction. These included 73 patients from the buparlisib plus paclitaxel arm and 71 from the paclitaxel plus placebo arm (**Figure 2**). Baseline characteristics of patients with evaluable FFPE samples for H&E analysis are presented in **Table 1**.

**Figure 2.**
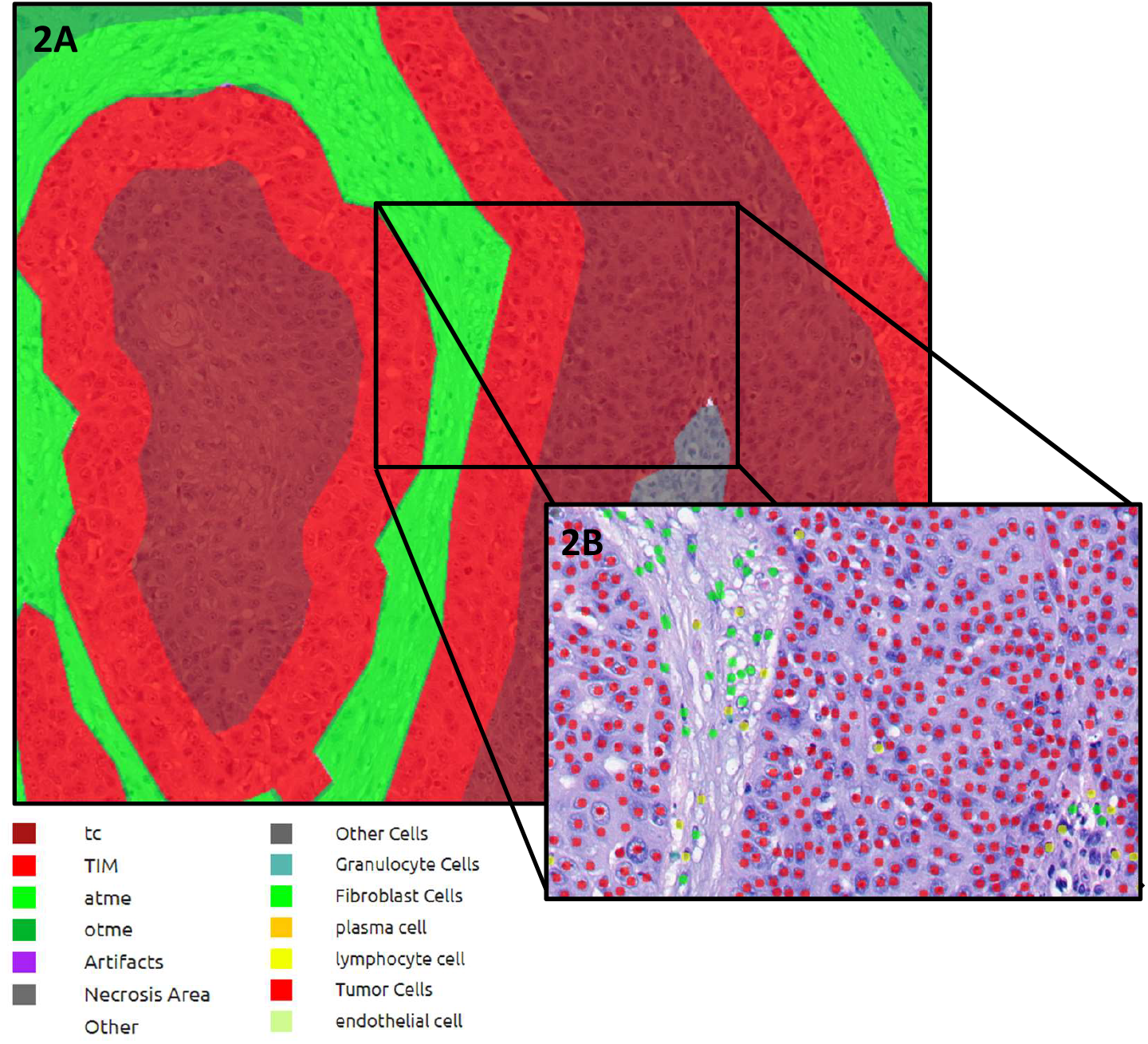
Spatial annotation of tumor and immune microenvironment regions and single-cell phenotypes in head and neck squamous cell carcinoma histopathology. Figure 2A: Representative hematoxylin and eosin (H&E) stained whole-slide image processed with automated spatial segmentation. Tumor and stromal regions are delineated based on proximity to the tumor-stroma interface. The tumor region includes the *tumor center* (TC) and *tumor invasive margin* (TIM), defined as the 60 µm internal band adjacent to the tumor boundary. The tumor microenvironment (TME) includes the *adjacent TME* (aTME), a 60 µm external band, and the *outer TME* (oTME) beyond that. Each zone is color-coded and overlaid for visualization. Figure 2B. High-resolution region showing overlay of cell-type annotations derived from a deep learning classifier. Individual cells are labeled by phenotype, including tumor cells (red), lymphocytes (green), fibroblasts (yellow), endothelial cells (blue), and others. Color legend indicating spatial compartments (top row) and single-cell phenotypes (bottom row). This integrative digital pathology framework enables spatial biomarker analysis across defined tumor and microenvironment zones. TC, tumor center; TIM, tumor invasive margin; aTME, adjacent tumor microenvironment; oTME, outer tumor microenvironment; TME, tumor microenvironment (aTME + oTME); Tumor area = TC + TIM.

### Deep learning model performance for tissue and cell classification

To support spatial biomarker extraction, we first evaluated the performance of the deep learning models for histologic region segmentation and single-cell classification. The histological area segmentation model showed strong concordance with expert annotations, with accuracy rates of 85.0% for tumor, 92.8% for stroma, 80.9% for necrosis, and 90.6% for nerve regions (**Figure 3A**). The cell classification model accurately identified major stromal and immune phenotypes, achieving the highest agreement for lymphocytes (83.5%), tumor cells (76.6%), fibroblasts (70.6%), and plasma cells (61.9%) (**Figure 3B**). Performance for granulocytes was moderate (70.7%), while endothelial cell classification was more limited (48.2%). All predictions were reviewed for biological plausibility and integrated into spatially contextualized cell maps, which served as the basis for subsequent biomarker extraction across TME compartments. While plasma cell misclassification with lymphocytes and moderate granulocyte accuracy were noted, these predictions were considered sufficient for exploratory analyses, though granulocyte detection from H&E will require additional validation in future studies.

**Figure 3.**
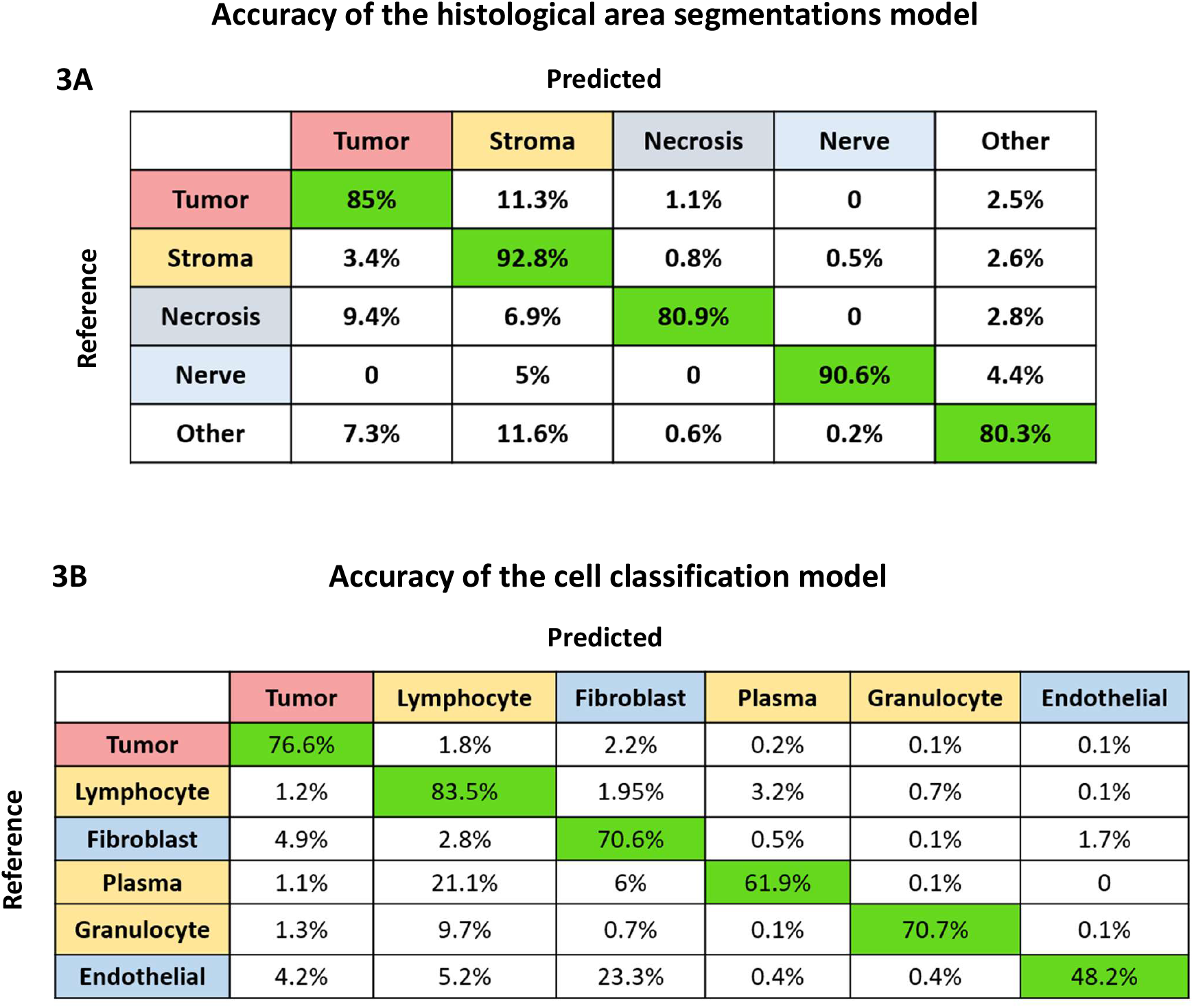
Performance metrics of deep learning models for area segmentation and single-cell classification in head and neck squamous cell carcinoma. Figure 3A. Area segmentation model performance showing percent agreement between predicted and reference labels for five histological regions: tumor, stroma, necrosis, nerve, and other. High concordance is observed for tumor (85.0%), stroma (92.8%), necrosis (80.9%), and nerve (90.6%). Figure 3B: Cell classification model performance with percent agreement across seven cell types: tumor cells, lymphocytes, fibroblasts, plasma cells, granulocytes, and endothelial cells. Classification accuracy is highest for tumor (76.6%), lymphocytes (83.5%), fibroblasts (70.6%), and plasma cells (61.9%), with moderate accuracy for granulocytes (70.7%) and endothelial cells (48.2%). Correct classifications are highlighted in green.

### Quantitative evaluation of tumor-infiltrating lymphocytes using H&E vs immunohistochemistry

The prognostic and predictive relevance of TILs was assessed using AI-based spatial quantification of H&E images compared to conventional IHC scoring. Among patients with high TIL density (≥10%), a significant treatment effect of buparlisib versus placebo was observed in patients with high TILs (HR□=□0.25; 95% CI, 0.01–0.64), whereas no between-arm difference was seen in patients with <10% TILs (HR□=□0.89; 95% CI, 0.60–1.34) (**Figure 4A**). Exploratorily, within the placebo arm, OS trended longer in patients with <10% TILs than in those with ≥10% TILs, whereas no clear within-arm difference was seen in the buparlisib arm; these within-arm high-vs-low comparisons were descriptive and not powered for inference. In contrast, IHC-based TIL scoring yielded a weaker association (HR□=□0.51 in the ≥10% group; 95% CI, 0.26–1.00; HR□=□0.83 in the <10% group; 95% CI, 0.52–1.33) (**Figure 4B**). In a secondary analysis of patients with baseline tumor samples evaluable for HPV status by p16 immunohistochemistry (n=70 buparlisib; n=73 placebo; see **Figure 1**), the proportion of HPV-positive tumors was higher among cases with ≥10% TILs than among those with <10% TILs (**Supplemental Figure 1**), irrespective of treatment arm or primary site.

**Figure 4:**
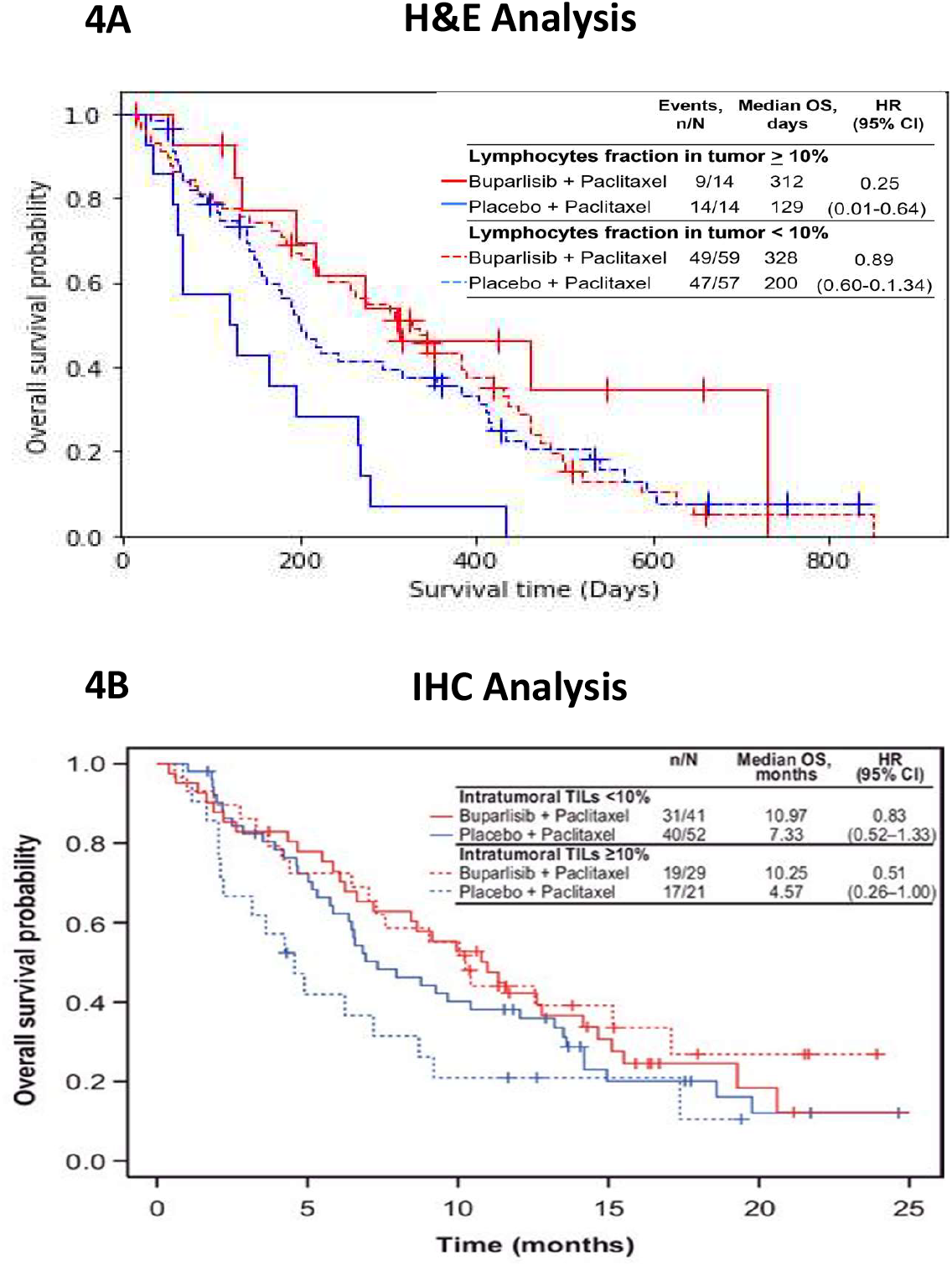
Tumor-infiltrating lymphocytes as a predictive biomarker of buparlisib response: comparative analysis using H&E-based spatial quantification and IHC-based scoring. Figure 4A. H&E-based lymphocyte fraction analysis: Kaplan-Meier curves of overall survival (OS) stratified by lymphocyte fraction in tumor regions as quantified from H&E-stained slides. Patients with ≥10% lymphocyte infiltration in the tumor area showed markedly improved OS when treated with buparlisib + paclitaxel compared to placebo (HR = 0.25; 95% CI, 0.01–0.64). In contrast, patients with <10% lymphocytes did not show a survival benefit (HR = 0.89; 95% CI, 0.60–1.34). Figure 4B. IHC-based tumor-infiltrating lymphocyte (TIL) analysis: Kaplan-Meier curves of OS based on IHC-assessed intratumoral TILs. Among patients with ≥10% TILs by IHC, buparlisib + paclitaxel conferred a survival benefit over placebo (HR = 0.51; 95% CI, 0.26–1.00), albeit with less discrimination than H&E-based analysis. In the <10% TILs subgroup, no significant difference in OS was observed between treatment arms (HR = 0.83; 95% CI, 0.52–1.33). Both H&E- and IHC-based TIL analyses suggest that high baseline lymphocytic infiltration in the tumor microenvironment may serve as a prognostic and potentially predictive biomarker for benefit from buparlisib in combination with paclitaxel. H&E-based spatial annotation demonstrated stronger hazard ratios and clearer OS separation than traditional IHC scoring.

### Tumor microenvironment heterogeneity

We next assessed the association between TME cellular heterogeneity and clinical outcomes using the Shannon entropy index derived from AI-based H&E analysis. Among patients with high TME heterogeneity (above median), a significant treatment effect of buparlisib versus placebo was observed in patients with high TME heterogeneity (HR□=□0.47; 95% CI, 0.27– 0.80; p□=□0.005), whereas no between-arm difference was seen in patients with low TME heterogeneity (**Figure 5A**). In contrast, no between-arm difference was observed in patients with **low** TME heterogeneity (HR□=□0.99; 95% CI, 0.60–1.65; p□=□0.99) (**Figure 5B**). These findings suggest that increased cellular and spatial complexity within the TME may enhance sensitivity to PI3K inhibition and serve as a predictive biomarker of buparlisib benefit, rather than reflecting general prognostic differences.

**Figure 5.**
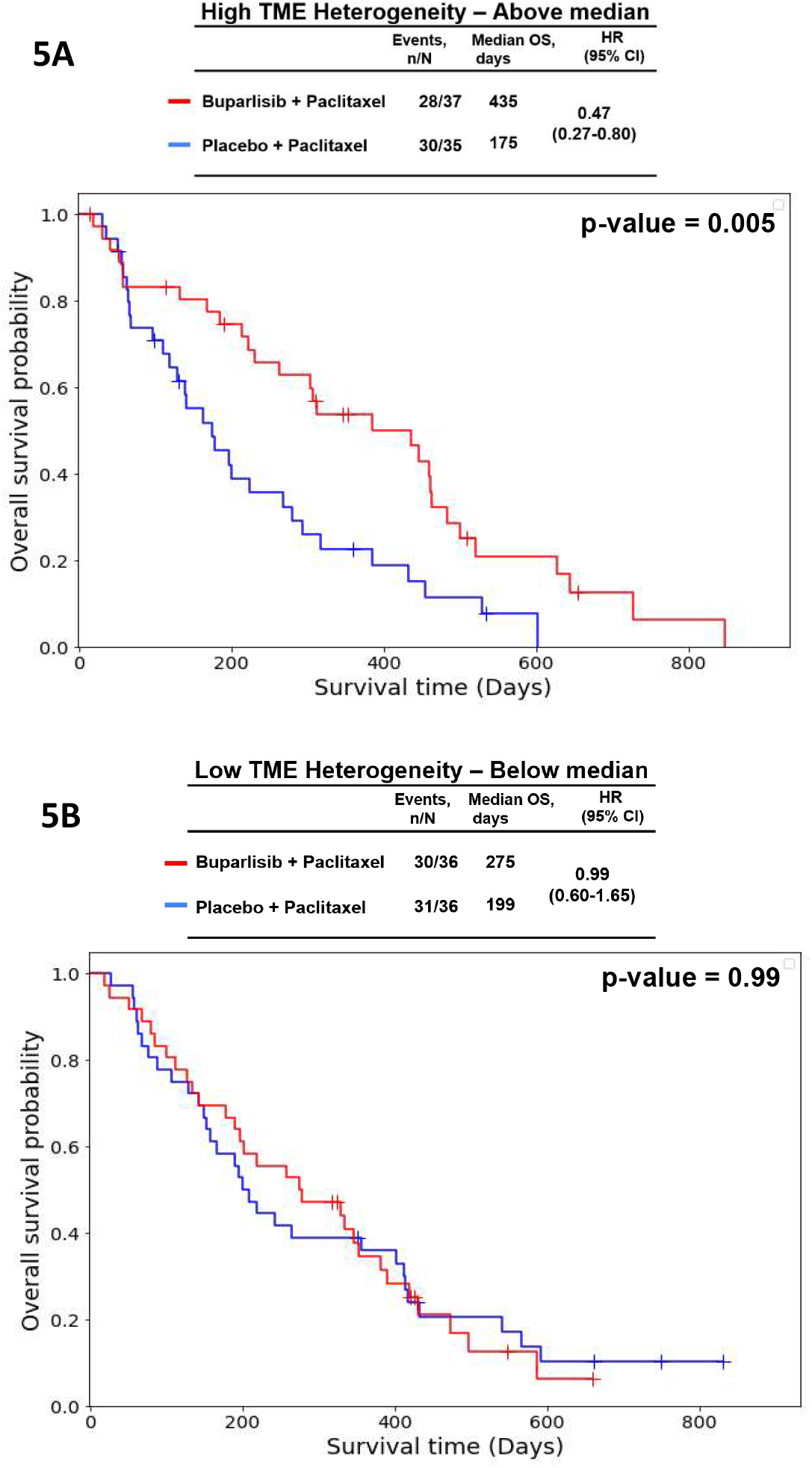
Tumor microenvironment cellular heterogeneity as a predictor of overall survival in buparlisib-treated patients. Figure 5A. Kaplan-Meier curves of overall survival (OS) stratified by tumor microenvironment (TME) cellular heterogeneity, derived from H&E-based digital pathology. High versus low TME heterogeneity was defined according to the median cutoff. In patients with high TME heterogeneity, treatment with buparlisib + paclitaxel was associated with significantly improved OS (HR = 0.47; 95% CI, 0.27–0.80, p = 0.005) compared to patients treated with placebo + paclitaxel. Figure 5B. No OS difference in patients with low TME heterogeneity was seen between the buparlisib + paclitaxel and the placebo + paclitaxel groups (HR = 0.99; 95% CI, 0.60–1.65, p = 0.99).

### Granulocyte enrichment in the tumor invasive margin

Among patients with high granulocyte infiltration in the tumor invasive margin (TIM), as quantified by AI-based H&E analysis, buparlisib was associated with significantly improved OS compared to placebo (HR = 0.51; 95% CI, 0.30–0.88; p = 0.014) (**Figure 6A**). In contrast, no between-arm difference was observed in patients with low granulocyte infiltration (HR = 0.93; 95% CI, 0.56–1.54; p = 0.78) (**Figure 6B**). In a complementary within-arm proximity analysis, a higher percentage of granulocytes in proximity to tumor cells was strongly associated with improved OS among patients receiving buparlisib + paclitaxel (HR = 0.32; p = 0.00006), consistent with a predictive rather than purely prognostic effect.

**Figure 6.**
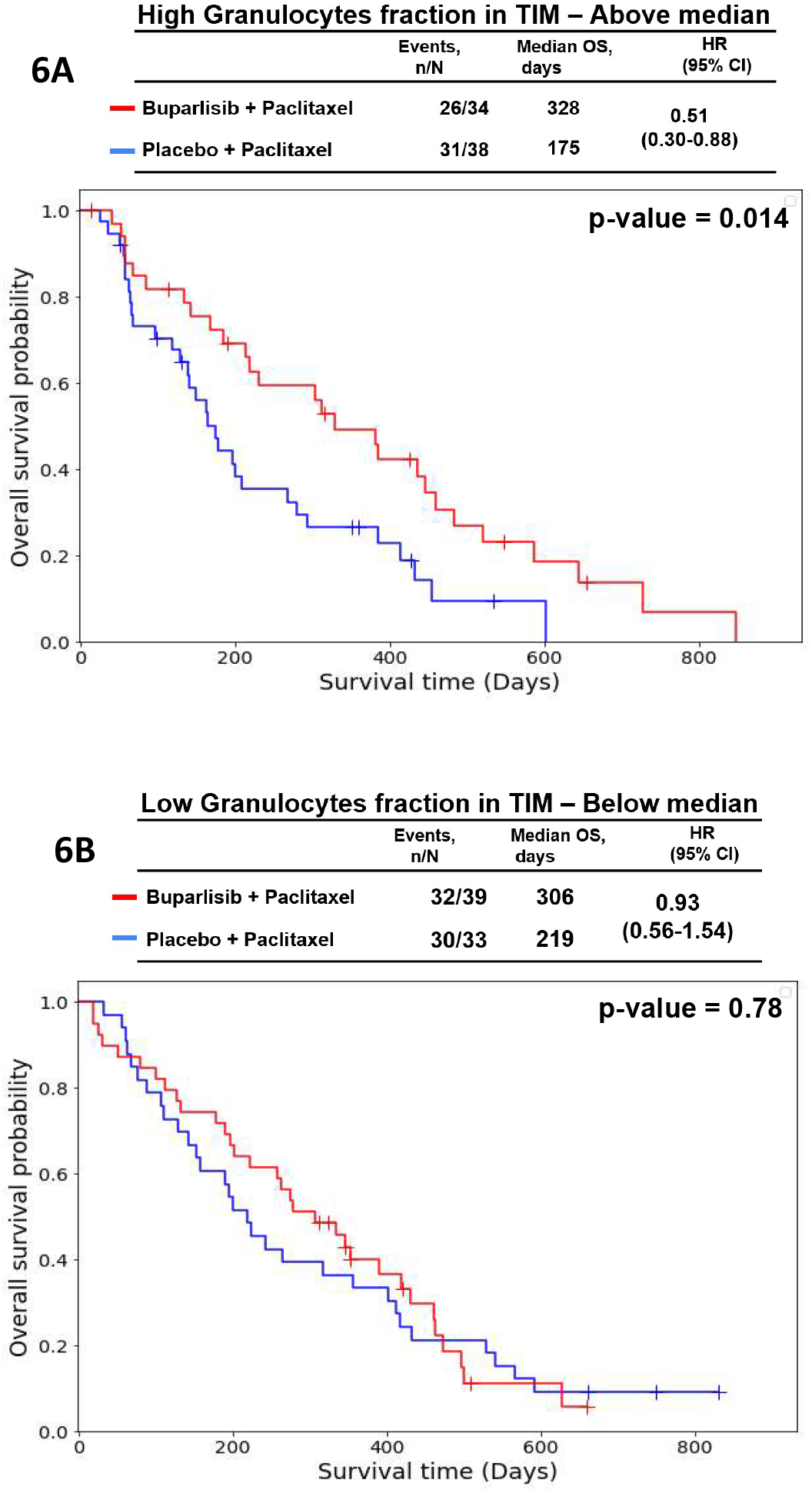
Granulocyte enrichment in the tumor invasive margin (TIM) as a predictor of overall survival in buparlisib-treated patients. Figure 6A. Kaplan-Meier curves of overall survival (OS) stratified by granulocyte fraction in the TIM based on H&E-derived digital pathology. High versus low granulocyte fraction was defined according to the median cutoff. In patients with high granulocyte fraction, treatment with buparlisib + paclitaxel was associated with significantly improved OS (HR = 0.51; 95% CI, 0.30–0.88, p=0.014) compared to patients treated with placebo + paclitaxel. Figure 6B. No OS difference in patients with low granulocyte fraction was seen between the buparlisib + paclitaxel and the placebo + paclitaxel groups (HR = 0.93; 95% CI, 0.56–1.54, p=0.78).

## Discussion

This study demonstrates that spatial biomarkers derived from AI-enabled analysis of H&E images can predict therapeutic benefit from buparlisib in R/M HNSCC. Specifically, elevated tumor-TILs, high tumor TME heterogeneity, and increased granulocyte infiltration in the tumor invasive margin (TIM) were each associated with improved treatment effect of buparlisib in OS – suggesting a treatment-specific predictive effect rather than a purely prognostic one.

These findings reinforce and extend prior BERIL-1 biomarker studies based on genomic and immunohistochemical profiling^8,9^, while demonstrating the enhanced discriminatory capacity of H&E-based spatial metrics. Notably, AI-derived TIL quantification produced stronger hazard ratios and clearer outcome separation than conventional IHC scoring, highlighting the potential of digital pathology to refine immune-related biomarkers. While the confidence interval for OS in high-TIL patients was wide (HR = 0.25; 95% CI, 0.01–0.64), the directionality and significance of the finding remain robust.

Similarly, high TME heterogeneity was associated with improved treatment effect of buparlisib versus placebo in OS (HR = 0.47; 95% CI, 0.27–0.80), underscoring the importance of spatial and cellular diversity in modulating response to PI3K inhibition. Elevated granulocyte infiltration in the TIM was also linked to improved buparlisib treatment effect (HR = 0.51; 95% CI, 0.30–0.88), suggesting that granulocytes at the tumor–stromal interface may exert an immunomodulatory role that synergizes with PI3K inhibition and chemotherapy. This potential interaction between innate immune infiltration and targeted PI3K blockade provides a mechanistic rationale for why these spatial biomarkers are particularly relevant in the buparlisib plus paclitaxel setting.

Although model performance was generally strong, the lower classification accuracy for endothelial cells (48.2%) reflects the challenge of rare or morphologically ambiguous cell types and indicates an area where further refinement would be beneficial.

This study also underscores the clinical utility of H&E-based spatial analysis in identifying candidates most likely to benefit from buparlisib plus paclitaxel. By capturing both established and spatially complex features of the TME, AI-driven models can detect immune infiltration, heterogeneity, and stromal interactions not easily discernible by conventional methods. These results provide mechanistic insights into buparlisib’s mode of action and inform patient selection strategies for future PI3K-targeted studies. Spatial biomarkers derived from H&E are cost-effective, interpretable, and scalable across pathology workflows – key advantages that support their ongoing validation in the BURAN phase 3 trial.

This study has several limitations. It was a retrospective, post hoc analysis of a single phase II trial with a moderate sample size (n = 144 evaluable slides), limiting power for subgroup and interaction effects. All biomarker analyses were exploratory, with nominal two-sided p-values and no adjustment for multiplicity, increasing the risk of type I error. While the deep learning pipeline performed well overall, detection accuracy for certain phenotypes (e.g., granulocytes, endothelial cells) was only moderate, and H&E-based classification remains inherently imperfect. No independent, blinded validation set or prospectively locked thresholds were used, and variability in slide quality and digitization could have introduced noise or bias. Clinical confounding cannot be excluded in this paclitaxel combination setting, and generalizability beyond BERIL-1 is uncertain. Accordingly, these results should be regarded as hypothesis-generating and require prospective validation. The ongoing BURAN phase III trial includes pre-specified biomarker definitions, centralized scanning, blinded algorithmic reads, and interaction testing to prospectively evaluate these spatial biomarkers.

## Conclusion

AI-based spatial analysis of routine H&E slides offers a scalable, non-destructive approach to identify predictive biomarkers of buparlisib efficacy in R/M HNSCC. These spatial features capture both established immune biomarkers, such as lymphocyte infiltration, and additional aspects of tumor–stromal architecture not detected by traditional assays. Their dual relevance underscores the biological plausibility of these findings in the context of PI3K inhibition plus chemotherapy and highlights their potential utility for patient selection. These findings support the integration of digital pathology into biomarker-guided trial design and underscore the relevance of immune and stromal architecture in modulating PI3K inhibitor response. Prospective validation is warranted to confirm their clinical utility in additional validation cohorts and guide personalized treatment strategies.

## Data Availability

The datasets generated and/or analyzed during the current study are available from the corresponding author upon reasonable request.

## Declarations

### Ethics approval

This study was conducted in accordance with Good Clinical Practice guidelines, local regulatory requirements, and the ethical principles of the Declaration of Helsinki. Ethics approval was obtained from the institutional review boards or ethics committees at all participating sites. A study steering committee provided oversight to ensure protocol compliance, and an independent data monitoring committee regularly reviewed safety data.

### Consent to participate

All participants provided written informed consent prior to study enrollment.

### Consent for publication

All authors have reviewed and approved the final version of this manuscript and consent to its submission for publication.

### Author contributions

**A.D**.: Conceptualization, Software, Formal analysis, Data Curation, Writing - Original, Draft, Writing - Review & Editing, Visualization;: **M.T.L**.: Conceptualization, Software, Formal analysis, Data Curation, Writing - Original, Draft, Writing - Review & Editing, Visualization; **J.L**.: Conceptualization, Methodology, Validation, Formal analysis, Resources, Writing - Review & Editing, Project administration; **O.M.N**.: Methodology, Formal analysis; **A.B**.: Methodology, Formal analysis; **A.L**.: Methodology, Formal analysis; **S.R.Z**.: Methodology, Formal analysis; **Z.R**.: Methodology, Formal analysis; **J.K.K**.: Methodology, Formal analysis; **A.G**.: Methodology, Formal analysis; **H.Y**.: Methodology, Formal analysis; **L.F.L**.: Conceptualization, Methodology, Investigation; **S.L**.: Conceptualization, Methodology; **A.T**.: Conceptualization, Methodology, Validation, Resources, Writing - Review & Editing, Project administration; **T.T**.: Conceptualization, Methodology, Resources, Project administration; **K.D**.: Conceptualization, Methodology, Resources, Project administration; **N.H**.: Conceptualization, Methodology, Resources, Project administration; **L.E.B**.: Conceptualization, Methodology, Resources, Project administration; **S.F**.: Conceptualization, Methodology, Investigation; **D.S**.: Conceptualization, Methodology, Validation, Formal analysis, Investigation, Writing - Review & Editing, Supervision, Project administration

## SUPPLEMENTAL FIGURES

**Supplemental Figure 1.**
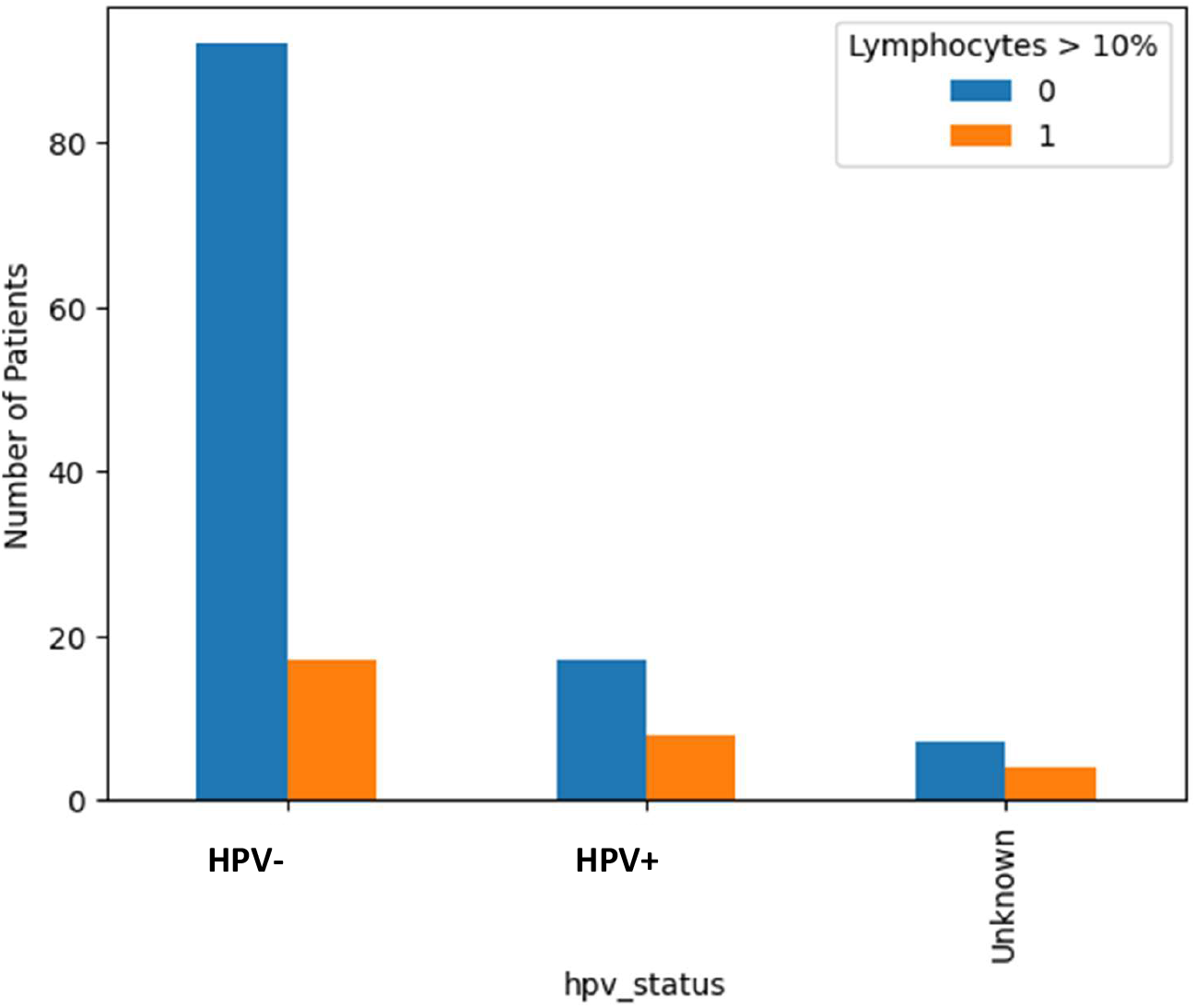
HPV status in patients with recurrent/metastatic head and neck squamous cell carcinoma was compared between those with ≥10% vs <10% tumor-infiltrating lymphocytes. HPV status was determined by p16 immunohistochemistry on baseline FFPE tumor samples, irrespective of tumor primary site.

